# Predicting Rejection Risk in Heart Transplantation: An Integrated Clinical–Histopathologic Framework for Personalized Post-Transplant Care

**DOI:** 10.1101/2025.09.05.25335209

**Authors:** Dale D. Kim, Anant Madabhushi, Kenneth B. Margulies, Eliot G. Peyster

**Affiliations:** Stanford University School of Medicine, Stanford, CA, USA; Department of Biomedical Engineering, Emory University, Atlanta, GA, USA; Division of Cardiovascular Medicine, University of Pennsylvania, Philadelphia, PA, USA; Wesley Research Institute, Brisbane, QLD, Australia

**Author notes:** **Corresponding Author:** Name: Eliot G. Peyster, Address: Level 8, East Wing, The Wesley Hospital, 451 Coronation Dr, Auchenflower QLD, Australia, 4066.

## Abstract

**Background:** Cardiac allograft rejection (CAR) remains the leading cause of early graft failure after heart transplantation (HT). Current diagnostics, including histologic grading of endomyocardial biopsy (EMB) and blood-based assays, lack accurate predictive power for future CAR risk. We developed a predictive model integrating routine clinical data with quantitative morphologic features extracted from routine EMBs to demonstrate the precision-medicine potential of mining existing data sources in post-HT care.

**Methods:** In a retrospective cohort of 484 HT recipients with 1,188 EMB encounters within 6 months post-transplant, we extracted 370 quantitative pathology features describing lymphocyte infiltration and stromal architecture from digitized H&E-stained slides. Longitudinal clinical data comprising 268 variables—including lab values, immunosuppression records, and prior rejection history—were aggregated per patient. Using the XGBoost algorithm with rigorous cross-validation, we compared models based on four different data sources: clinical-only, morphology-only, cross-sectional-only, and fully integrated longitudinal data. The top predictors informed the derivation of a simplified Integrated Rejection Risk Index (IRRI), which relies on just 4 clinical and 4 morphology risk facts. Model performance was evaluated by AUROC, AUPRC, and time-to-event hazard ratios.

**Results:** The fully integrated longitudinal model achieved superior predictive accuracy (AUROC 0.86, AUPRC 0.74). IRRI stratified patients into risk categories with distinct future CAR hazards: high-risk patients showed a markedly increased CAR risk (HR=6.15, 95% CI: 4.17–9.09), while low-risk patients had significantly reduced risk (HR=0.52, 95% CI: 0.33–0.84). This performance exceeded models based on just cross-sectional or single-domain data, demonstrating the value of multi-modal, temporal data integration.

**Conclusions:** By integrating longitudinal clinical and biopsy morphologic features, IRRI provides a scalable, interpretable tool for proactive CAR risk assessment. This precision-based approach could support risk-adaptive surveillance and immunosuppression management strategies, offering a promising pathway toward safer, more personalized post-HT care with the potential to reduce unnecessary procedures and improve outcomes.

**Clinical Perspective:** ***What is new?***

- Current tools for cardiac allograft monitoring detect rejection only after it occurs and are not designed to forecast future risk. This leads to missed opportunities for early intervention, avoidable patient injury, unnecessary testing, and inefficiencies in care.
- We developed a machine learning–based risk index that integrates clinical features, quantitative biopsy morphology, and longitudinal temporal trends to create a robust predictive framework.
- The Integrated Rejection Risk Index (IRRI) provides highly accurate prediction of future allograft rejection, identifying both high- and low-risk patients up to 90 days in advance – a capability entirely absent from current transplant management.

***What are the clinical implications?***

- Integrating quantitative histopathology with clinical data provides a more precise, individualized estimate of rejection risk in heart transplant recipients.
- This framework has the potential to guide post-transplant surveillance intensity, immunosuppressive management, and patient counseling.
- Automated biopsy analysis could be incorporated into digital pathology workflows, enabling scalable, multicenter application in real-world transplant care.

## Introduction

Heart transplantation (HT) is the treatment of choice for end-stage heart failure, with more than 6000 transplants performed worldwide in 2024.^1^ While HT is the most effective and durable therapy for patients with end-stage HF,^2^ transplanted hearts do not last forever. The recipient immune response is a persistent threat to allograft health, necessitating vigilant surveillance for early CAR detection and lifelong immunosuppression (IS) management to mitigate the risk of cardiac allograft rejection (CAR).^3–5^

The care HT patients receive is highly regimented, with the schedule for CAR surveillance testing and IS drug de-escalation determined by local institutional protocols. Within each institution, protocols are uniformly applied to the vast majority of HT recipients. Historically, EMB with histologic grading had been the sole diagnostic standard for CAR, and has been recommended by HT guidelines since 1990 despite concerns about inter-grader reliability and accuracy.^6–8^ More recently, ‘liquid biopsy’ methods using plasma-based cell-free DNA (cfDNA) and gene expression profiling (GEP) have been developed as less-invasive CAR screening alternatives for select patients, with positive results triggering confirmatory EMB.^9–13^ Regardless of approach, it is typical for an HT recipient to undergo 12 or more scheduled surveillance CAR tests and 12 or more scheduled changes in IS regimens during their 1^st^ year post-transplant alone.^14^

Due to “one-size-fits-all” protocols, patients at low risk for allograft complications undergo more diagnostic testing and receive more aggressive IS treatment than necessary.^15^ Over-aggressive IS increases the risk of infections, drug toxicity, and future malignancy. Excessive diagnostic testing increases both risk and cost. The prevalence of ‘false-positive’ cfDNA results still necessitate frequent EMBs and further invasive monitoring.^12^ And, though generally safe, EMBs still introduce risk, with procedural complications ranging from mild (e.g. access site hematoma, tricuspid regurgitation, and transient bundle branch block/arrhythmias) to severe (e.g. right ventricular perforation and cardiac tamponade).^16–18^ Rigid, uniform, and reactive protocols pose an even greater threat to patients at high-risk for future allograft complications who may experience inadequate surveillance and premature IS weaning. Furthermore, the extra resources required to manage these complications represents a form of potentially preventable waste in post-HT care.^19,20^ Ultimately, the frequency of CAR surveillance and the protocolized approach to IS weaning reflect a lack of tools for reliable, proactive, and individualized CAR risk assessment. While emerging benchmarks have improved the diagnosis of existing rejection events, neither the established EMB histologic grading criteria nor the more recently introduced cfDNA approaches are designed to predict rejection events weeks or months in the future. Reliable predictions are the basis for prospective risk stratification, which in turn is the basis for establishing personalized approaches to IS weaning and surveillance allograft schedules. Risk prediction tools have been utilized in clinical medicine for decades to improve the quality of physician decisions when presented with complex, competing-risk scenarios. While there have been limited studies in HT analyzing trends between certain clinical biomarkers and acute^21^ or chronic CAR,^22^ the evidence for robust predictive performance is lacking and these methods have not been adopted clinically. More recently, efforts to utilize computational analysis of digitized EMB slides have yielded some promising results for improving the diagnostic value of EMB,^23–26^ but have not yet been utilized to predict future CAR events. In this study, we generate an integrated clinical and morphologic model for prediction of future acute and subacute CAR after HT. Furthermore, we present a risk stratification approach for determining an individual’s likelihood of developing CAR. Notably, we identify distinct high and low risk groups in acute and subacute time windows to guide clinical decision-making surrounding IS adjustments and the need for invasive EMBs, providing a framework for precise, patient-centered care in HT management.

## Methods

### Study Cohort and Design

The goal of this research was to generate decision support models to identify patients with different CAR risk profiles who might benefit from more personalized post-HT management. We envision these models being used during early post-HT outpatient visits, when patients undergo routine testing such as bloodwork and EMBs. The study cohort was derived from patient records and archived histology samples at the University of Pennsylvania (UPenn) between 2007 and 2021, aiming to identify individuals who did vs. did not experience acute CAR during the first post-HT year.

For each patient, International Society for Heart and Lung Transplantation (ISHLT) histologic grades were extracted from all available EMB reports and binarized into “low-grade” CAR (ISHLT 0R or 1R) and “high-grade” CAR (ISHLT 2R or 3R, or any positive humoral rejection grade, pAMR > 0). In parallel, each EMB was labeled as either “clinically silent” or “clinically evident” rejection based on previously validated criteria for allograft injury.^23^ To assign these clinical severity labels, structured clinical metadata within ±7 days of each EMB—including symptoms, physical exam findings, lab tests, echocardiography, ECG, and hemodynamics—were reviewed (**Table S1).**

Digitized H&E-stained slides were obtained from four previously published cohorts,23– 26 and patients were included if they had at least one digitized EMB within the first 6 months post-HT. Patients were categorized as “Future Rejection” (FR) if they experienced either high-grade or clinically evident rejection within that 6-month period, or “No Rejection” (NR) if they did not. The final study cohort included 484 patients: 134 FR (320 digitized EMBs) and 350 NR (868 digitized EMBs). Among FR patients, EMBs were collected a mean of 39.75 (SD ±29.9) and median of 29.0 days before rejection.

### Clinical Dataset for Developing a Clinical Risk Model of Future CAR

Additional clinical data were extracted for each HT patient, including serial recipient medical history, medications, laboratory results, and echocardiograms. Donor data included medical history, home medications, labs during the procurement admission, HLA typing, cause of death, organ ischemic time, and recipient PRA and HLA match/mismatch.

### Quantitative Morphology Dataset for Developing a Morphologic Risk Model of Future CAR

Quantitative morphology data were extracted from digitized H&E EMBs using previously validated automated digital pathology pipelines.^24,26,27^ Analyses focused on two major histologic domains: lymphocytic immune infiltrates and stromal architecture. Immune-related features captured the size, density, and spatial organization of lymphocyte aggregates, while stromal features described fiber size and shape, orientation, accumulation, and spatial relationships to adjacent cardiomyocytes. In total, 370 features were generated per biopsy (154 immune-related and 210 stromal-related), with summary statistics aggregated to create EMB-level feature vectors representing the immune–stromal microenvironment. Full image-processing and feature-extraction procedures are detailed in the Supplementary Methods. **Figure 1** shows visual examples of both lymphocyte and stromal pipeline segmentation.

**Figure 1:**
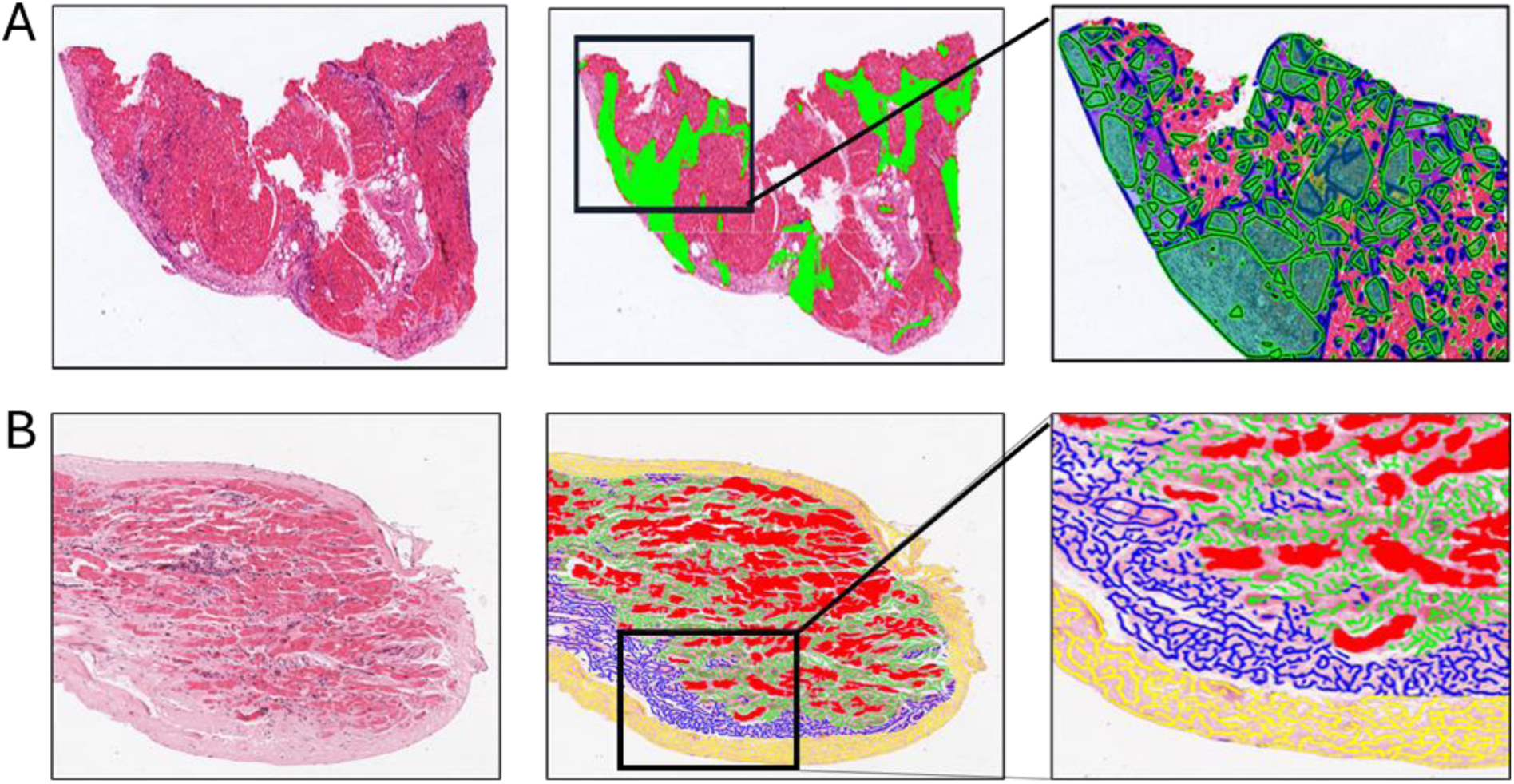
Quantitative morphology feature extraction examples. Digitized H&E whole-slide-images are analyzed by the lymphocyte feature pipeline (**A**) which identifies and measures lymphocyte colonies and foci and quantifies their relationship to surrounding myocyte structures, and by the stromal feature pipeline (**B**) which identifies stromal fibers in the digitized H&E slides, separating them into endocardial stroma, interstitial stroma (e.g. closely bounding cardiomyocytes), and stromal ‘islands’ which are macrostromal domains which are less closely associated with cardiomyocytes. The stromal fibers are then measured by area, orientation, and other geometrics and relational properties.

### Inclusion and Exclusion

To align with the goal of predicting rejection in an outpatient surveillance setting—and minimize early post-operative confounders—histology slides and clinical data from days 0–20 post-transplant were excluded. Only patients with ≥90% lab data completeness were included. For eligible patients with some missing data, Multiple Imputation by Chained Equations (MICE)^28^ was performed using the miceforest package (v6.0.3) in Python.

### Dataset Preparation and Statistical Analysis

Each outpatient visit with a digitized EMB was considered an ‘encounter’ for assessing future CAR risk. All clinical, demographic, and quantitative histology features were concatenated into a single matrix, yielding 1,188 encounters from 484 patients and 1,361 variables per encounter (**Table S2**). A consort diagram of the final cohort is shown in **Figure 2**. For each encounter, a comprehensive feature set integrated clinical, demographic, and morphologic data, including cross-sectional (e.g., day-of-visit), baseline (e.g., averaged first-month values), and longitudinal variables (e.g., change-over-time) (**Figure S1**). Full details on dataset and variable creation are described in the Supplemental Methods.

**Figure 2.**
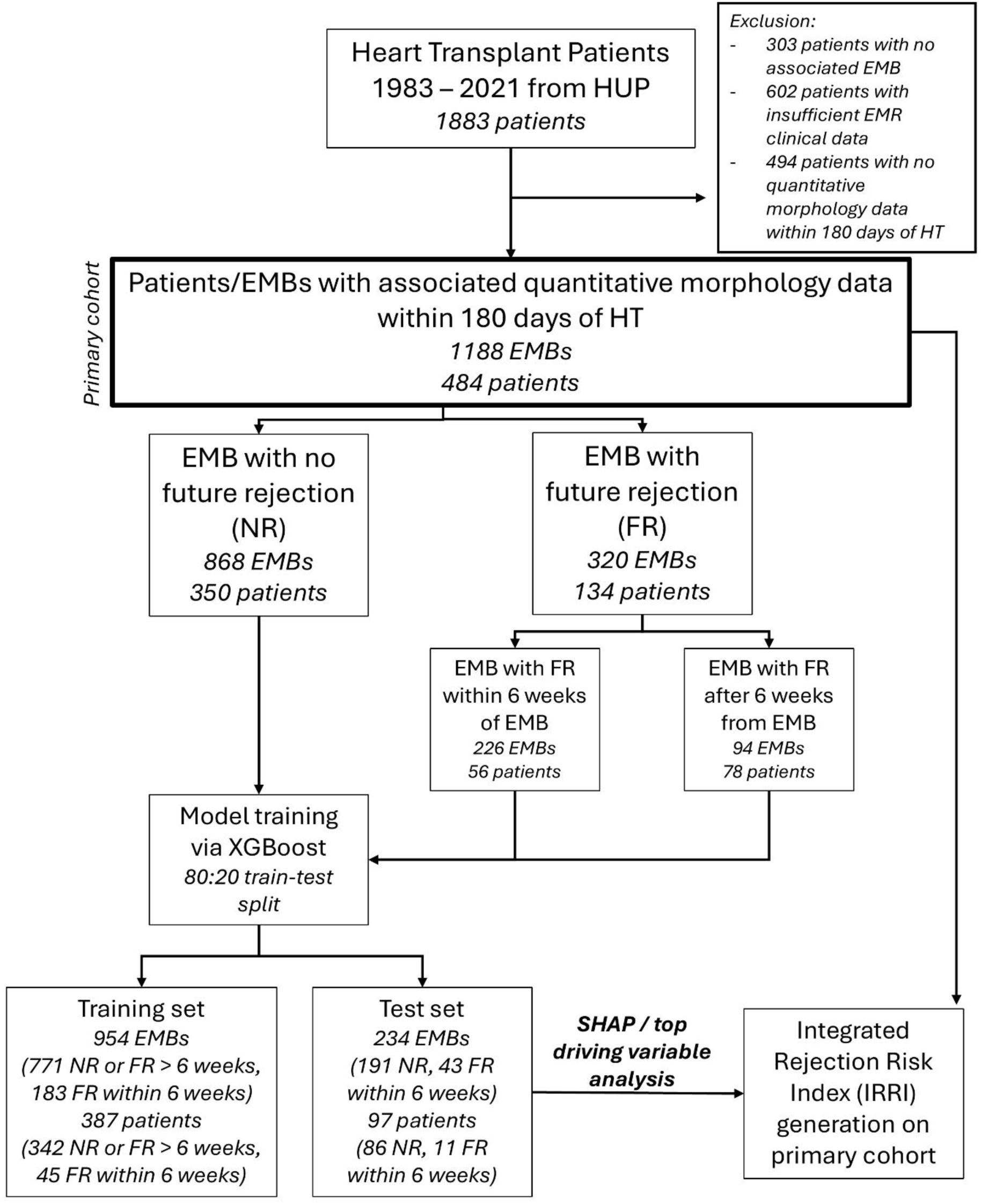
Consort diagram of patient cohort. Inclusion criteria for the primary cohort were patients with associated digitized EMBs and documented laboratory and medication data.

### Modeling Strategy

Models were generated to test three hypotheses: **1)** a Bayesian approach incorporating prior data and longitudinal trends would outperform cross-sectional data alone, **2)** adding quantitative morphologic data would enhance predictive power over clinical factors and rejection grade alone, and **3)** predictive information could be distilled into a small set of user-friendly risk factors identifying distinct patient-level CAR trajectories. Predictive modeling used XGBoost (v2.0.0) logistic classifiers, assigning ‘FR’ or ‘NR’ labels to each encounter. Data were split 80:20 into train/test sets by patient ID to avoid leakage, with 10-fold cross-validation for training. Final performance was assessed using area under the receiver operator curve (AUC), area under the precision-recall curve (PRC), and macro F1 score on the hold-out test set. Top predictors from the best model were extracted using SHAP feature selection (v0.43.0) to support interpretability and spatial effects,^29,30^ and to aid selection of a small number of discrete risk factors for potential clinical use. These were entered into a Cox proportional-hazards model to assess time-to-first-CAR-event during the first 6 months post-HT. Comparative trajectory performance between models was assessed with log-rank testing, hazard ratio (HR), and 95% confidence interval (CI).

## RESULTS

### Recipient and Donor Cohort Characteristics

Recipient and donor demographics and characteristics stratified by FR vs. NR status are described in **Table 1**. The final cohort consisted of N=484 patients with associated digitized EMBs, laboratory data, and medications data from within 180 days of their heart transplant, with N=134 FR patients and N=350 NR patients. Of the examined features, there were no statistically significant differences between the FR and NR cohorts for recipient or donor characteristics.

**Table 1.**
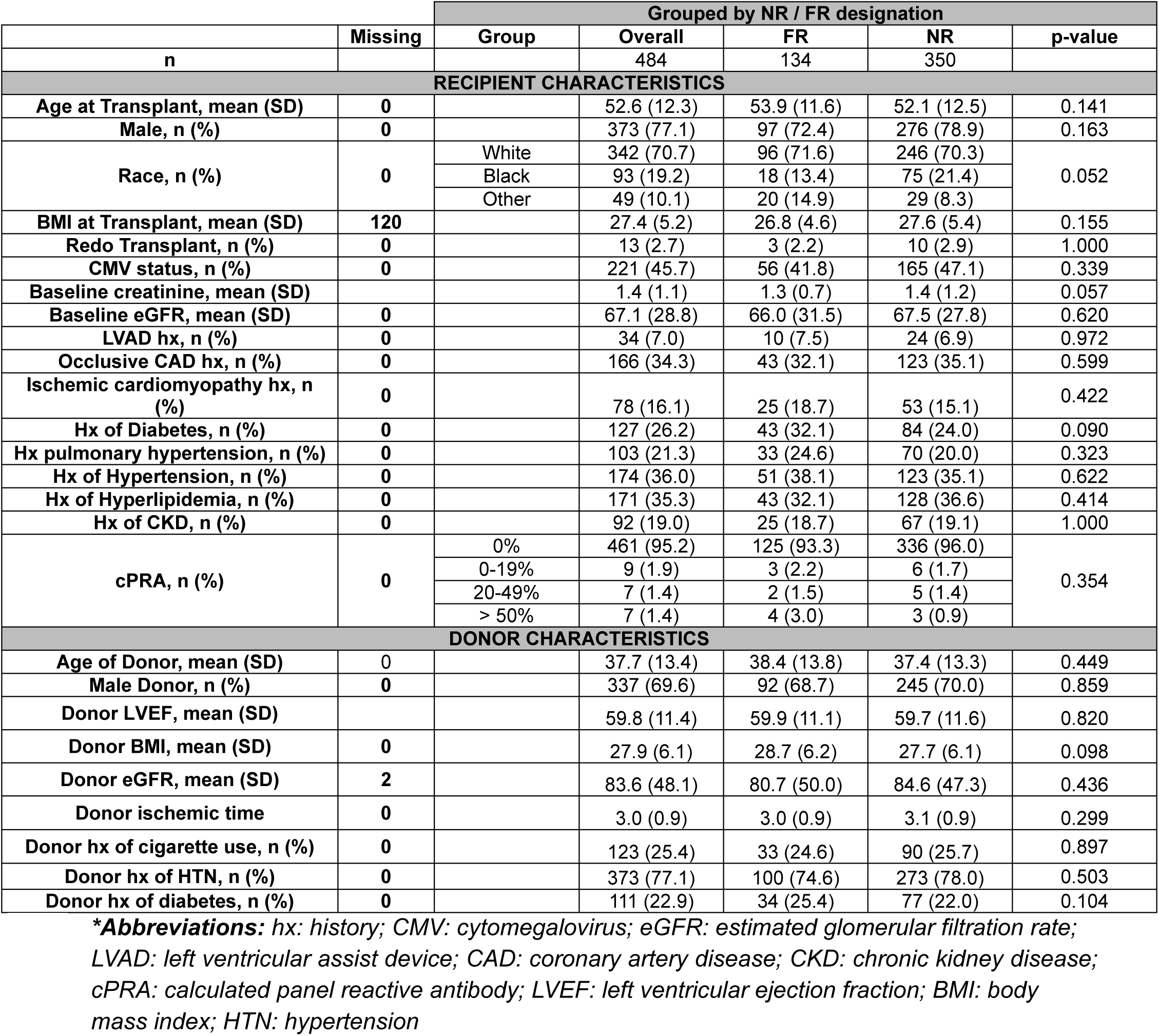
Descriptive statistics of recipients and donor baseline characteristics, stratified by whether the recipient is a “future rejector” (FR) or “never rejector” (NR).

### Predictive Modeling

#### Longitudinal Clinical Data Enhances Predictive Performance Over Cross-Sectional Approaches

Although physiologic and pathologic processes evolve continuously over time, clinical assessments are often limited to discrete, cross-sectional evaluations. Because laboratory, medication, and histopathology data are collected serially in HT patients, modern computational methods offer an opportunity to integrate and analyze this longitudinal information in a manner which better captures evolving pathologic processes. In our first experiment, we hypothesized that incorporating longitudinal trends in diagnostic and demographic data would significantly improve the prediction of CAR compared to relying on cross-sectional data alone.

Modeling performance was evaluated on the held-out test set for comparison. Using only routinely available, cross-sectional clinical data (n=185 variables) from study outpatient encounters, an XGBoost model achieved an AUROC of 0.62 and an AUPRC of 0.28 for predicting CAR within 6-weeks of the encounter, with a macro F1 score of 0.57, sensitivity of 0.53, and specificity of 0.70 (**Figure 3A–B**, **Table 2**). When longitudinal variables – such as percent change from first and most recent outpatient encounter, and change-over-time metrics – are integrated into the feature space (n=268 variables total), predictive performance improved substantially, with an AUROC of 0.77 (p=0.007 compared to cross-sectional only) and an AUPRC of 0.54 (p=0.0038 compared to cross-sectional only) with a macro F1 score of 0.70, sensitivity of 0.47, and specificity of 0.91 (**Figure 3A–D**, **Table 2**). These results highlight the added value of modeling dynamic patient trajectories in post-transplant care.

**Figure 3.**
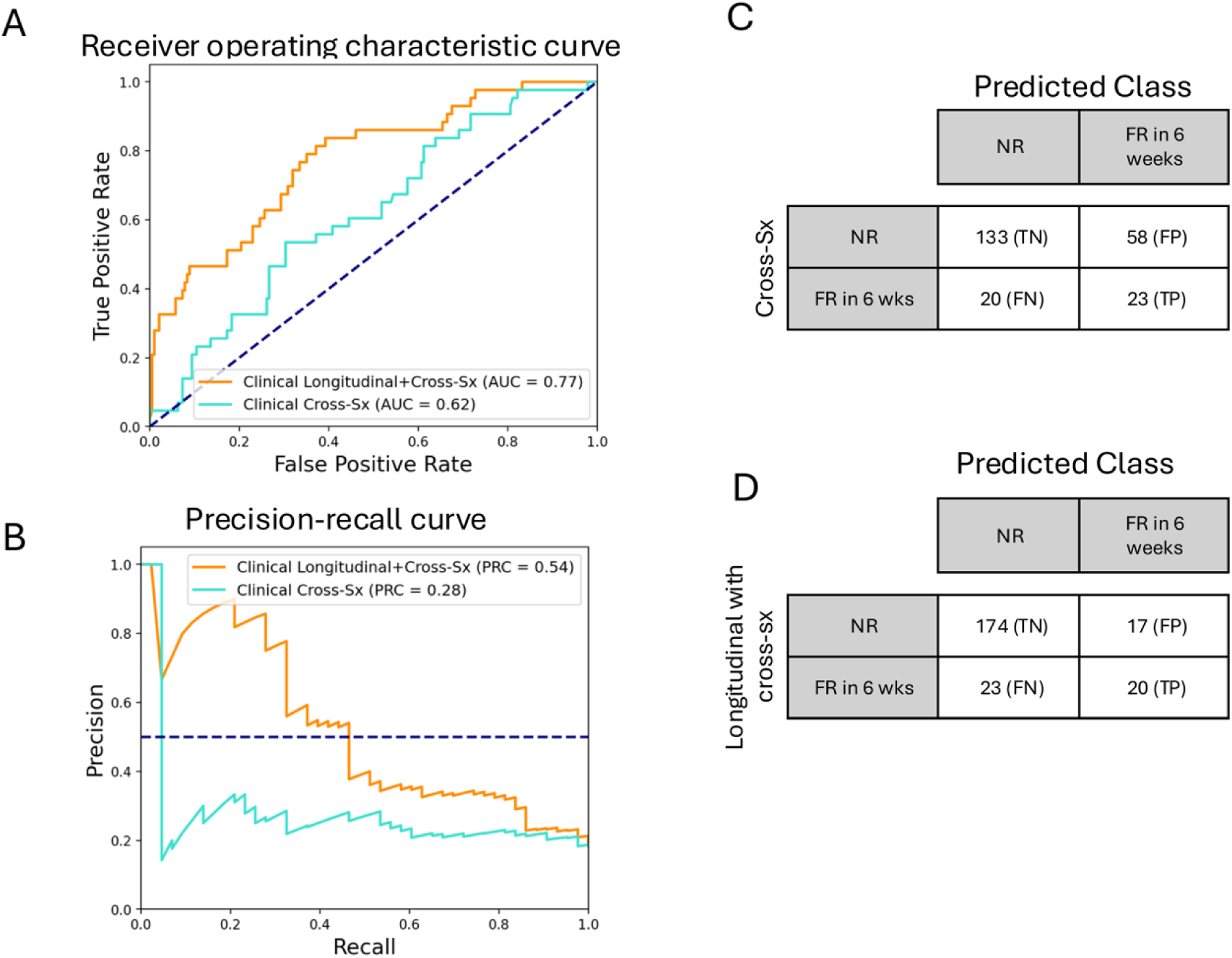
Performance of gradient-boosted models with stratified 10-fold cross-validation for cross-sectional-only vs. longitudinal and cross-sectional combined clinical variables in predicting future CAR within the first 180 days post-HT. **A)** AUROC and **B)** AUPRC curves for cross-sectional-only (blue) and longitudinal/cross-sectional combined (orange) feature spaces. Confusion matrices for **C)** cross-sectional-only predictors and **D)** longitudinal/cross-sectional combined predictors are shown.

**Table 2.** Performance reporting of models predicting rejection in different time windows and with different methods. Bootstrapping with N=1000 samples was used for statistical comparison between curves.

| Time window | Test Set N | # Pos. (%) | Feature Space | AUROC | AUPRC | Macro F1 | Sens. | Spec. | PPV | NPV | p-value |
| --- | --- | --- | --- | --- | --- | --- | --- | --- | --- | --- | --- |
| CLINICAL VARIABLES ONLY |  |  |  |  |  |  |  |  |  |  |  |
| 0-180d | 234 | 43 (18.4) | Cross-Sx only | 0.62 | 0.28 | 0.57 | 0.53 | 0.70 | 0.28 | 0.87 | 0.007 (ROC)<br>0.0038 (PRC) |
|  |  |  | Cross-Sx and longitudinal combined | 0.77 | 0.54 | 0.70 | 0.47 | 0.91 | 0.54 | 0.88 |  |
| INTEGRATED CLINICAL-MORPHOLOGIC MODEL |  |  |  |  |  |  |  |  |  |  |  |
| 0-180d | 234 | 43 (18.4) | Cross-Sx only | 0.72 | 0.47 | 0.66 | 0.40 | 0.90 | 0.47 | 0.87 | 0.013 (ROC)<br>0.011 (PRC) |
|  |  |  | Cross-Sx and longitudinal combined | 0.86 | 0.74 | 0.83 | 0.72 | 0.94 | 0.74 | 0.94 |  |
| CROSS-SECTIONAL AND LONGITUDINAL VARIABLES COMBINED |  |  |  |  |  |  |  |  |  |  |  |
| 0-180d | 234 | 43 (18.4) | Clinical | 0.77 | 0.54 | 0.70 | 0.47 | 0.91 | 0.54 | 0.88 | 0.057 (ROC)<br>0.029 (PRC) |
|  |  |  | Integrated | 0.86 | 0.74 | 0.83 | 0.72 | 0.94 | 0.74 | 0.94 |  |

**\*Abbreviations:** cross-sx: cross-sectional; AUROC: area under the receiver operating characteristic curve; AUPRC: area under the precision-recall curve; Sens.: sensitivity; Spec.: specificity; PPV: positive predictive value; NPV: negative predictive value

#### Multi-Modality Integration Further Improves Prediction

Building on prior evidence of histopathologic utility in HT rejection assessment,^23–26^ we next evaluated whether integrating quantitative morphologic features extracted from the endomyocardial biopsy (EMB) performed during each study encounter, together with corresponding clinical data, could improve predictive performance.

The integrated clinical-morphologic model outperformed the clinical-only model in predicting CAR within 6-weeks of the encounter, achieving an AUROC of 0.86 (p=0.057 compared to clinical-only) and an AUPRC of 0.74 (p=0.029 compared to clinical-only) with a macro F1 score of 0.83, sensitivity of 0.72, and specificity of 0.94 (**Figure 4A–B**, **Table 2**). Across all tested models, this integrated approach yielded the highest performance metrics (**Figure 4C**). Best-performing model parameters and training performance are provided in **Table S3.**

**Figure 4.**
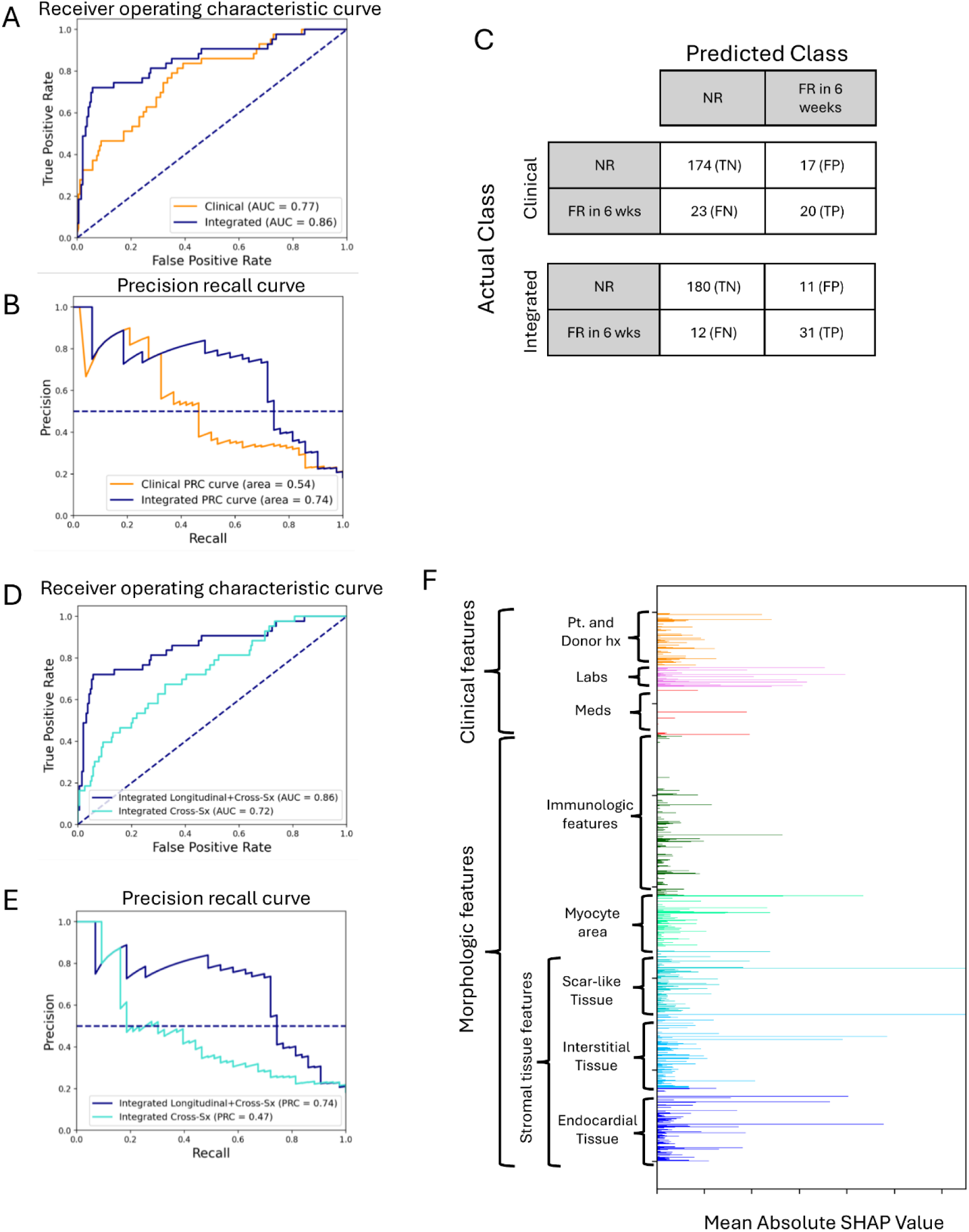
Performance and feature space of clinical-only and morphology-integrated models for predicting 6-week future CAR within the first 180 days post-HT. **A)** receiver-operating curve, **B**) precision-recall curve, and **C)** confusion matrix for clinical-only (orange) vs. morphology-integrated (dark blue) models. **D)** receiver-operating curve and **E**) precision-recall curve for cross-sectional-only integrated (light blue) and longitudinal/cross-sectional morphology integrated (dark blue) models. **F)** Global feature space overview of main contributors to the best-in-class prediction model using SHAP analysis.

As with clinical data alone, incorporating longitudinal features into the integrated dataset substantially improved performance for predicting future CAR events compared to a cross-sectional-only dataset (**Figure 4D–E**, **Table 2**).

To provide insights on the relative importance of different classes of variables for predicting future CAR, we performed SHAP analysis of the longitudinal integrated model (**Figure 4F**). In general, we observed that morphologic features tended to contribute larger magnitudes to the model, with significant contributions coming from stromal tissue features, while myocyte area ratio and immunologic indices had smaller but still significant contributions. Among clinical features, dosage changes or weaning of immunosuppressive medications (e.g. discontinuation of mycophenolate mofetil) showed the greatest impact, while laboratory and demographic variables contributed more broadly but with lower magnitude.

Together, these findings demonstrate that combining longitudinal and multi-modal data streams yields superior predictive performance, supporting a multifactorial, precision-medicine approach to post-transplant rejection risk modeling.

#### Translating Signal into Practice: Development of a Clinically Actionable Risk Stratification Approach for Future CAR Prediction

While the XGBoost model in the previous section provides a powerful method for assessing the feasibility of predicting future CAR, the complex and high-dimensional feature space of gradient-boosted models are a barrier to interpretation and real-world application. To facilitate clinical translation, we developed the Integrated Rejection Risk Index (IRRI), a simplified composite scoring system which leverages top features from our XGboost method to create a user-friendly framework for CAR risk stratification.

To develop IRRI, we first identified the most influential features in the XGBoost model using SHAP values. From these, we prioritized variables that were both biologically meaningful and routinely available in the electronic medical record (and via automated morphology analysis). Selected features were grouped into two domains—Clinical and Morphologic—to reflect distinct but complementary dimensions of CAR prediction.

Figure 5A summarizes the four Clinical and four Morphologic criteria comprising IRRI. Clinical items cover renal function, protein/liver abnormalities, prior EMB grade, and recent immunologic risk factors; morphologic items capture endocardial thickening, interstitial expansion, stromal:myocyte ratio, and lymphocyte infiltration. Each domain yields a 0–4 composite score. Further specifics on generating morphologic criteria are described in the Supplemental Methods and **Table S4**.

**Figure 5.**
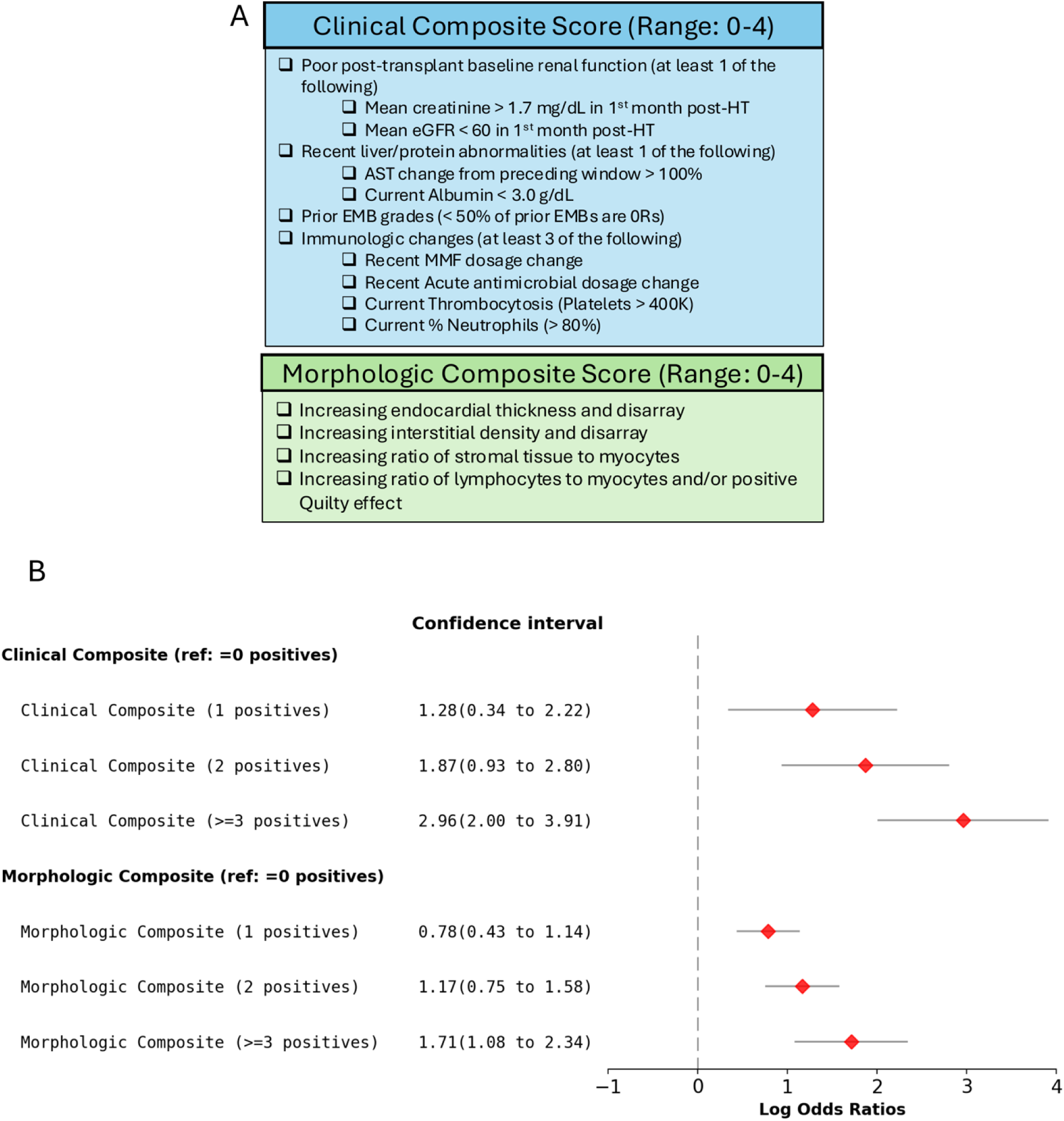
Development of an end-user-friendly approach to stratifying low- and high-risk clinical and morphologic patients. **A)** Model-derived criteria for clinical and morphologic composite scores. **B)** Log-odds scores of developing any future CAR within the first 180 days post-HT for different risk groups, compared to a reference of either 0 clinical composite positives or 0 morphologic composite positives, respectively.

#### Assessing predictive potential of IRRI clinical and morphologic risk criteria

As shown in Figure 5B, logistic regression analysis demonstrated that patients with any clinical or morphologic IRRI risk criteria had significantly increased odds of future CAR than patients without criteria. Specifically, patients with high clinical risk (a composite score of 3 or more) had a log-odds ratio of 2.96 (95% CI: 2.00-3.91) compared to the no-risk group, reflecting nearly a 20-fold CAR risk increase in odds. High morphologic risk, defined similarly, was associated with a log-odds ratio of 1.71 (95% CI: 1.08-2.34), representing a 5.5-fold increase in odds.

#### Leveraging IRRI risk criteria to identify distinct future CAR risk trajectories

Combining both clinical and morphologic IRRI criteria, we identified three distinct HT patient risk trajectories for experiencing CAR within 90 days of a patient encounter, as defined below:

- **Low Risk** (N=127): ≤1 total risk criteria
- **Intermediate Risk** (N=313): ≤2 clinical and ≤2 morphologic risk criteria
- **High Risk** (N=44): high risk in one domain (score ≥3) plus at least intermediate risk in the other.

Kaplan-Meier time-to-event analysis revealed clear separation in outcomes across these risk strata (Figure 6A**–C**). Encounters labeled high-risk by integrated IRRI had a 75.6% CAR rate within 90 days—significantly higher than clinical-only (43.7%, p<0.001) or morphologic-only (39.0%, p<0.001) high-risk groups. Similarly, the low integrated IRRI risk group showed enhanced negative predictive value: only 10.7% of these encounters were followed by CAR during the subsequent 90 days, significantly fewer than those labeled low-risk by clinical (17.7%, *p*=0.024) or morphologic criteria (26.1%, *p*<0.001).

**Figure 6.**
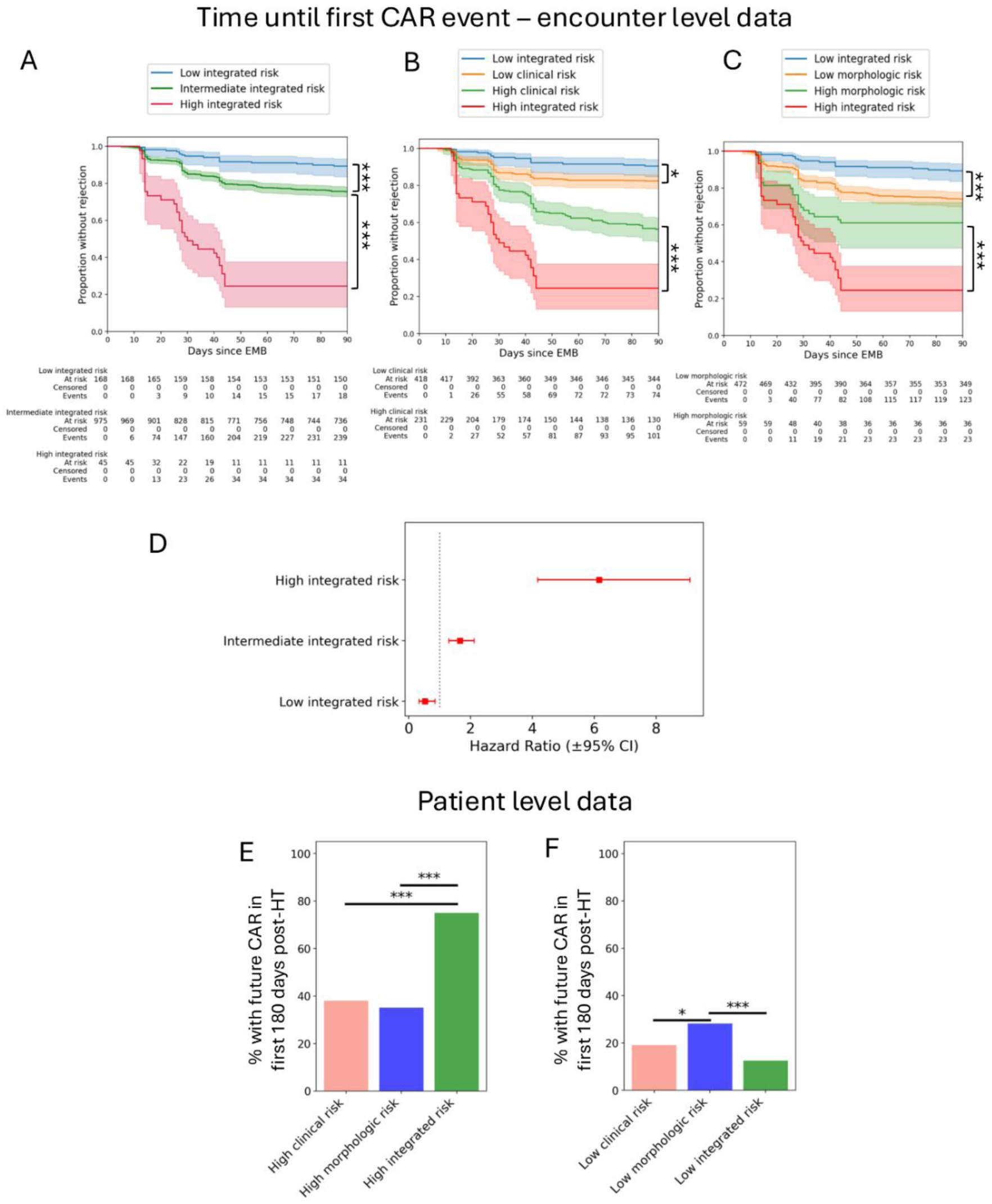
Time-to-rejection trajectory comparisons for clinical-only, morphologic-only, and integrated composite score risk groups. Encounter-level trajectory comparison between **A)** low, intermediate, and high integrated risk groups, **B)** low- and high-risk clinical-only and integrated risk groups, and **C)** low- and high-risk morphologic-only and integrated risk groups. **D)** Cox-regression hazard ratios of future CAR after an encounter between low, intermediate, and high integrated risk groups. **E and F)** Patient-level stratification based on the highest-risk encounter during the study period. **(E)** Patients with high integrated IRRI risk had significantly higher rates of future CAR compared to clinical- or morphologic-only high-risk groups. **(F)** Patients with low integrated IRRI risk had the lowest observed rates of CAR within 180 days post-HT. Kaplan-Meier curves compared for statistical significance using log-rank testing and proportions compared using Z-test for proportions (**\*** p<.05, **\*\*** p<.01, **\*\*\*** p<.001).

Cox regression confirmed these trends: high integrated IRRI risk encounters had a hazard ratio (HR) of 6.15 (95% CI: 4.17–9.09) for future CAR, compared to low-risk encounters. Intermediate-risk encounters carried a modest increase in risk (HR=1.65, 95% CI: 1.29–2.11), while low-risk encounters demonstrated a protective association (HR=0.52, 95% CI: 0.33–0.84) (Figure 6D).

To further validate these findings at the patient level, we analyzed outcomes based on each patient’s single highest-risk encounter during the 180-day post-HT period (Figure 6E-F). Those meeting high integrated IRRI criteria at any point within 180 days post-HT had a 75.0% probability of future CAR—significantly higher than high clinical-only (38.0%, p<0.001) or high morphologic-only (35.2%, p<0.001) groups. Low integrated-risk patients had a 12.6% CAR rate, lower than low clinical risk (19.0%, p=0.15) and significantly lower than low morphologic risk (28.2%, p<0.001).

Together, these encounter-level and patient-level analyses underscore the added value of integrating clinical and histomorphologic domains in a unified risk framework. The IRRI scoring system provides a simple yet powerful tool for identifying patients at heightened short-term CAR risk and guiding post-transplant surveillance and treatment intensity accordingly.

## DISCUSSION

CAR remains a persistent threat to both short- and long-term graft survival after HT In this study, we demonstrate the feasibility of prospective CAR prediction using machine learning (ML) applied to longitudinal clinical and morphologic data. By integrating serially collected laboratory results, immunosuppression records, and computational features extracted from EMBs, we developed a robust predictive model capable of anticipating CAR events up to six weeks in advance. Building on this, we translated model outputs into a practical, interpretable tool—the IRRI—to stratify patients by future CAR risk with high predictive accuracy. These findings have direct implications for enabling precision surveillance and individualized IS management in post-transplant care.

### Temporal Dynamics and Multi-Domain Integration Drive Predictive Performance

A central insight from this work is that CAR risk is better captured by temporal trends than by static clinical snapshots. Rejection often emerges gradually, preceded by subtle, cumulative shifts in lab values, medication doses, and tissue morphology. Incorporating these dynamic features into our models yielded substantial performance gains, improving performance across all data domains and ultimately reaching AUROC of 0.86 with full multi-modal integration. These findings highlight the value of modeling patient trajectories over time, rather than relying on single-time-point measurements.

Importantly, embedding biopsy morphometrics alongside clinical trends yielded superior predictive value compared to either domain alone. While previous studies examining the utility of clinical biomarkers for CAR prediction have been published,^31,32^ our findings emphasize the need for a robust framework consisting of multiple, diverse features, including parameters distinct from standard ISHLT grading criteria. Features such as endocardial stromal thickening, interstitial expansion, and increased stromal-to-myocyte ratio, and subtle differences in overall lymphocyte counts were among the strongest independent predictors of future rejection. These results reinforce that EMBs remain a critical source of biologic insight—not only diagnostically, but prognostically—when enhanced by computational methods that quantify histologic features in a consistent and reproducible fashion.

This study builds upon and extends a growing body of digital pathology work in cardiac transplantation. Prior studies have demonstrated that computational morphometrics could match expert pathology grading,^24^ could outperform conventional grading for assessing rejection severity,^26^ and could be leveraged to provide predictive insights into long-term outcomes such as graft dysfunction and allograft vasculopathy.^23,25,27^ Our current work represents a next phase in this trajectory—moving from retrospective analysis and diagnosis toward prospective forecasting and clinical decision support.

### Morphologic Predictors Reflect Signals of Subclinical Immune Injury

Notably, the top-performing morphologic predictors in our model likely reflect underlying biology associated with early immune activation and tissue remodeling. For example, increased stromal-to-myocyte ratio and interstitial expansion may represent extracellular matrix accumulation driven by low-grade inflammation. Endocardial thickening may reflect expansion of the subendocardial matrix, edema, or Quilty lesions—each being histologic manifestations of chronic alloimmune stimulation. Similarly, increased lymphocytic infiltration in the absence of meeting formal rejection criteria may signal ongoing but subclinical immune surveillance, and adds to prior work which highlighted the long-term prognostic implications of recurrent ‘low-grade’ rejection events.^27^

These observations suggest that computational histology offers a window into the biologic continuum of rejection, capturing tissue-level signs of inflammation, damage, and repair before overt rejection is detected. This capability complements and enhances the utility of traditional EMB grading, offering the potential to shift transplant care from reactive to proactive.

### From Model Performance to Practical Translation: The Role of IRRI

Past studies utilizing ML approaches to transplant rejection generally focus on model performance and optimization.^33–37^ In the present study, we combine best-in-class model outputs with clinical domain knowledge to bridge the gap between high-dimensional ML model outputs and bedside application. The result of this effort was IRRI—a simplified scoring system derived from eight key predictors (four clinical, four morphologic) identified using SHAP-informed feature attribution. IRRI classifies patients into low-, intermediate-, and high-risk categories for future CAR, providing easily understood and potentially actionable, patient-level risk stratification.

Despite its simplicity relative to the XGBoost ML model from which it is derived, IRRI preserves strong predictive power, with a positive predictive value of 75.6% in the high-risk group and a negative predictive value of 89.3% in the low-risk group. These results suggest that with further refinement and validation, IRRI could serve as a practical tool to guide personalization of CAR surveillance and IS decisions. For instance, low-risk patients might safely defer some routine EMB and cfDNA testing visits, reducing procedural burden, anxiety, and cost. Conversely, high-risk patients may benefit from delayed IS tapering, closer follow-up, or even preemptive therapeutic intervention before histologic signs of rejection emerge.

This risk-adaptive approach supports a move toward value-based transplant care— aligning resource use with individual risk and reducing both under- and over-treatment. It also has the potential to improve interpretation of emerging non-invasive tests. While cfDNA assays offer a sensitive method for detecting graft injury, they suffer from false positives that can prompt unnecessary biopsies. IRRI, on the other hand, offers a high specificity and precision method for predicting future graft injury, with decent but not superior sensitivity. Thus, IRRI in combination with cfDNA could offer a powerful integration of complementary modalities of CAR assessment that warrants further investigation.

Prospective studies will be needed to define IRRI’s role in clinical workflows— particularly in combination with cfDNA—and to determine whether IRRI-guided surveillance protocols can safely reduce CAR testing burden while maintaining or even improving outcomes for high-risk patients.

### Translating Routine Data into Actionable Risk: Scalable Implementation Potential of IRRI

A key advantage of IRRI is its reliance on data already collected in routine post-HT care: standard laboratory tests, medication records, and H&E-stained EMBs. Unlike omics platforms or specialized assays, IRRI can be implemented without additional patient burden, major infrastructure changes, or ‘send-out’ tests which take days to result. Digital clinical data are routinely stored in structured EHRs, and histology slides are easily digitized in most clinical pathology labs. As such, IRRI can be deployed as a software-based decision support tool – either locally or via cloud platforms – at relatively low cost and with the potential to provide rapid assessments to providers.

This approach builds on the principle that most of the tools needed to implement precision transplant medicine already exist. The challenge is not in data acquisition but in how existing data are synthesized, interpreted, and translated into action. IRRI addresses this challenge by leveraging computational techniques to convert fragmented longitudinal data into a coherent, actionable score.

### Limitations and Future Directions

Several limitations warrant acknowledgment. First, this was a single-center study, and findings may reflect local practice patterns, demographic factors, or biopsy protocols. While the quantitative morphology pipeline was externally validated previously,^24^ external validation across diverse institutions will be necessary to establish generalizability of IRRI. Second, real-world implementation of IRRI will require seamless integration with clinical workflows, including automated data extraction from the EHR and biopsy digitization pipelines. Third, while our use of data binning facilitated temporal modeling, it may obscure short-term fluctuations relevant to acute rejection events. Future iterations could explore higher-resolution modeling using real-time data streams.

Additionally, while IRRI focuses on clinical and morphologic domains, incorporating other rejection surveillance modalities like cfDNA, echocardiogram, or ECG could further refine and contextualize risk assessments. Prospective interventional trials are also needed to determine whether IRRI-guided care improves outcomes, reduces unnecessary testing, or safely supports more individualized IS strategies.

## Conclusion

This study demonstrates that machine learning applied to routine post-transplant data can generate high-performing, interpretable models for future rejection prediction. By integrating longitudinal clinical features with quantitative biopsy morphometrics, and distilling these into the IRRI scoring system, we offer a practical framework for personalized CAR surveillance and IS management.

Our findings support a shift away from rigid, generalized post-HT protocols toward a more targeted, risk-adaptive strategy, rooted in the specific risk profiles of individual patients. With future validation, enhancements and clinical integration, IRRI has the potential to improve outcomes, reduce harm, and reshape the standard of care in heart transplantation.

## Data Availability

Raw de-identified data will be made broadly available upon publication of this manuscript.

## Acknowledgements

The authors would like to thank the Penn Pathology Clinical Services Center (Li-Ping Wang, Jeffrey Gordon and Amy Ziober) for their efforts in support of this research. We would also like to acknowledge the computational efforts of Sara Arabyarmohammadi and Cai Yuan.

## Supplemental Material

Supplemental Methods

Tables S1-S4

Table S2 (.xlsx file, large dataset)

Figures S1-S2

## Non-standard Abbreviations and Acronyms

CAR: cardiac allograft rejection
cfDNA: cell-free DNA
GEP: gene expression profiling
IS: immunosuppressant

